# A Tool for Low-Cost, Quantitative Assessment of Shoulder Function Using Machine Learning

**DOI:** 10.1101/2023.04.14.23288613

**Authors:** David M. Darevsky, Daniel A. Hu, Francisco A. Gomez, Michael R. Davies, Xuhui Liu, Brian T. Feeley

## Abstract

Tears within the stabilizing muscles of the shoulder, known as the rotator cuff (RC), are the most common cause of shoulder pain—often presenting in older patients and requiring expensive, advanced imaging for diagnosis^1–4^. Despite the high prevalence of RC tears within the elderly population, there are no accessible and low-cost methods to assess shoulder function which can eschew the barrier of an in-person physical exam or imaging study. Here we show that a simple string pulling behavior task, where subjects pull a string using hand-over-hand motions, provides a reliable readout of shoulder health across animals and humans. We find that both mice and humans with RC tears exhibit decreased movement amplitude, prolonged movement time, and quantitative changes in waveform shape during string pulling task performance. In rodents, we further note the degradation of low dimensional, temporally coordinated movements after injury. Furthermore, a predictive model built on our biomarker ensemble succeeds in classifying human patients as having a RC tear with >90% accuracy. Our results demonstrate how a combined framework bridging task kinematics, machine learning, and algorithmic assessment of movement quality enables future development of smartphone-based, at-home diagnostic tests for shoulder injury.

## Introduction

Chronic pain and musculoskeletal (MSK) injuries are the most common cause of disability in the USA^5^. However, prior to referring patients for either operative or non-operative interventions, patients are first triaged based on their exact MSK pathology, which not only requires an in-person physical exam but also often demands costly and resource-intensive imaging, such as an MRI scan. Unfortunately, the elderly, patients in rural communities, and groups historically underrepresented in medicine face the greatest demographic and socioeconomic barriers^6,7^ to seeking in-person care, thus accounting for significant deficits in quality-of-life^8^. These groups are therefore hindered in their ability to rehabilitate from MSK injuries by dual obstacles: (1) the diagnosis of their MSK injury and (2) regular follow-up to track recovery. One method to improve healthcare access within underserved communities is developing a technological framework to both remotely track joint health and evaluate recovery from MSK pathology using inexpensive tools such as motion capture and algorithmic assessment of movement quality.

To develop such technology, we focused on the shoulder joint, which is one of the most often injured joints in the human body in part because its extensive range of motion (ROM) relies on complex muscular and soft tissue supports. Movement across the shoulder’s ROM is accomplished through the synergistic action of two muscle groups: those that move the shoulder through its ROM and those that stabilize the shoulder joint during movement^4^. These latter muscles, collectively known as the “rotator cuff” (RC), are commonly injured resulting in >30% population prevalence of symptomatic RC pain with advancing age^1,2,8^. RC tears progressively worsen across time, which leads to considerable physical disability and limitations in activities of daily living due to pain, range of motion loss, and deficits in neuromuscular control of the shoulder^1,2,8^.

Animal models allow for precise control over both the temporal specificity and the extent of shoulder injury. Thus, we used a mouse model of RC injury known to recapitulate the histopathological features of chronic human RC tears^9,10^ to develop a machine learning-driven pipeline for quantifying motion quality as it relates to shoulder function. In contrast to other preclinical studies of rodent shoulder kinematics, which rely on quadruped gait analysis that is not translatable to bipedal humans, we introduced a novel preclinical model of shoulder function that builds upon the string pulling behavior task—a bimanual, oscillatory forelimb movement (Fig. 1) where mice rope-in a string like a sailor pulling cables on a sail ship^11^. Critically, string pulling task performance is highly conserved across rodents and humans thus raising the intriguing possibility of applying our algorithmic pipeline for assessing shoulder health in rodents directly to humans^12^.

**Figure 1.**
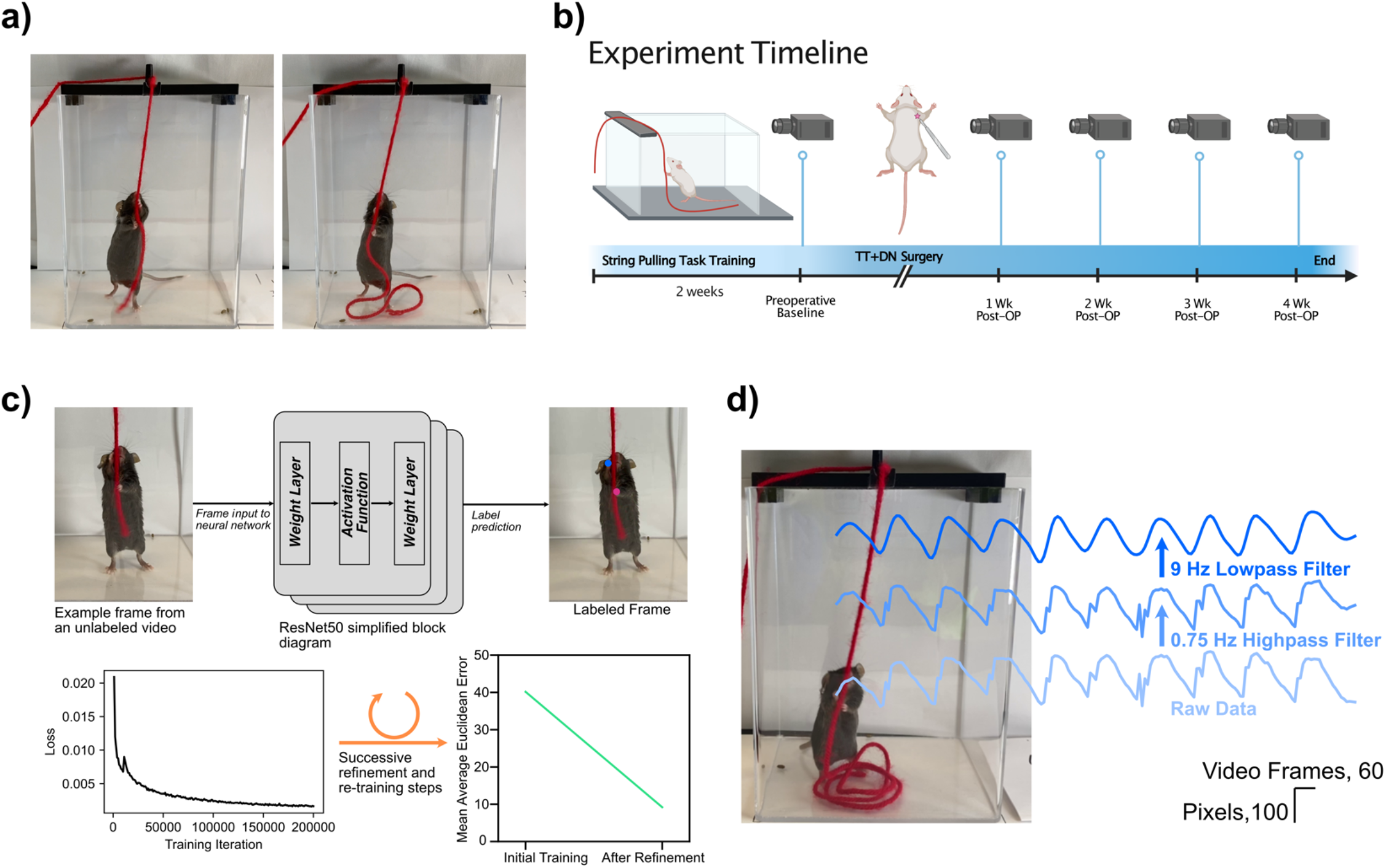
Overview of string pulling behavior and video pre-processing. **a)** String pulling behavior. Mice pull on a piece of string, held in a reproducible location across trials with a 3D printed string holder, using hand-over-hand motions similar to a sailor pulling cables on a sail ship. **b)** Experiment timeline. Mice (n=12) were pretrained three times per week on the task before a preinjury video recording was completed. Mice were then given a rotator cuff injury via surgical transection of the supraspinatus and infraspinatus tendons along with transection suprascapular nerve. Half the mice received immediate repair of the injured tendons while the other half received no repair. Mice were allowed to recover for one week prior to the commencement of weekly recordings. **c)** *Top row*: schematic showing labeling of video frames using a pretrained ResNet50 deep convolutional neural network. *Bottom row, left*: Example decay in root mean square error loss across neural network training. *Bottom row, right*: After two refinement steps, average Euclidean error on a held out test set drops from 40.16 to 9.21 pixels on a held out test set. **d)** Kinematic trajectory filtering. Example of worst case scenario jitter in trajectory labels (data shown for right hand in the Y (vertical)-axis). Highpass followed by lowpass Butterworth filtering eliminates low frequency drift and high frequency jitter in trajectory labeling, respectively.

Previous methods for automated assessment of shoulder function in humans relied on expensive marker- or sensor-based techniques^13–18^. Instead, we capitalize on recent advances in machine learning to implement low-cost markerless motion capture of string pulling behavior videos^19^. We first developed our video-based biomarkers of shoulder function using a rodent model for RC tears and then validated concordance of our preclinical biomarkers in human patients with rotator cuff pathology previously diagnosed using MRI scans. Here we found a striking cross-species concordance in biomarkers associated with RC injury. These results provide preliminary validation of our methodology for assessing movement quality using an inexpensive assay (see Supp. Table 1 for list of material costs) that can be readily deployed either in community health clinics, or even in a patient’s home, as either a screening test for shoulder pathology or as a tool to track recovery of kinematics following shoulder injury or surgery.

## Results

### Behavioral apparatus and data collection methodology

To standardize recordings of string pulling behavior in mice, a 3D printed string holder was placed over the behavior box to maintain a consistent position of the string across animals and behavioral trials (Fig. 1a). The position of the video camera relative to the behavior box was also standardized across recordings by using an alignment jig, and all videos were recorded at 59.64 frames per second (FPS).

Mice were first acclimated to a plexiglass behavior box for two days prior to the start of string pulling training. Following acclimation, mice (n=12) received two weeks of string pulling training conducted three times per week (Fig. 1b). A Cheerio attached to the end of the string served as a reward for trials in which the mouse pulled the string all the way into the behavior box. The initial training period was followed by a preoperative baseline behavioral recording where each mouse was recorded pulling a 0.75 m long string for a total of two trials (∼20-30 sec of data per animal; see Supplementary Movie 1 for representative baseline string pulling behavior). Animals then underwent surgery with combined supraspinatus (SS)/infraspinatus (IS) tendon transection and denervation of the right shoulder; half of the animals (n=6 mice) received immediate repair of the SS/IS tendons^9,10^. After animals recovered for one week, string pulling behavior was recorded for an additional four weeks.

After completion of data collection, we used DeepLabCut, a package for training deep convolutional neural networks for automated image segmentation^19^, to extract locations and labels of the right and left hands (Fig. 1c, top). In brief, 50 video frames were extracted from each recorded video and labeled by manual curation. These videos were then used for supervised transfer learning of a ResNet50 deep convolutional neural network (CNN) that was pre-trained on ImageNet (Fig. 1c, middle top). Feed forward inference was then performed on all video frames in the dataset to automatically label the right and left hands (Fig. 1c, right top). After an initial round of CNN training and inference, we extracted frames where the labels for at least one of the hands jumped by a Euclidean distance of 20 or more pixels. These frames were relabeled and the CNN retrained; this refinement step reduced the mean Euclidean error in label prediction from 40.16 to 9.21 pixels on a randomly selected 5% set of held-out test images (Fig. 1c, bottom). On rare occasions, a mouse’s hands would be occluded by the string causing brief oscillations in the labeling (see Fig. 1d, “Raw Data” trace for a worst-case scenario example). We thus filtered the hand trajectories first with a 0.75 Hz 1^st^-order Butterworth high pass filter (to remove any contributions from low frequency postural changes), followed by 9Hz 3^rd^-order Butterworth low pass filter (to remove any oscillations in hand labeling secondary to hand occlusion). Filter frequencies and orders were selected to minimize distortion of the kinematic trajectory.

### Post-processing of string pulling trajectories

Figure 2a demonstrates the overlay of 10 cycles of string pulling behavior for an example mouse prior to injury (right arm in blue, left arm in red). We found that the oscillatory nature of the behavior resulted in movement trajectories that occurred predominately in the vertical Y-axis. Temporally unrolling the Y-axis trace revealed that reach epochs evolved faster than pull epochs as the latter required mice to apply downward force as they advance the string (Fig. 2b, every other reach/pull epoch labeled for visualization purposes only). For each reach and pull epoch, we calculated its duration (in number of video frames divided by frame rate) and amplitude (measured in number of pixels). At baseline, Pearson correlations of the filtered kinematics traces for the right vs left hand in the X and Y axes revealed a high correlation in side-to-side (X-axis) movements of the arms (Median r= 0.7660, Q1 = 0.6245, Q3 = 0.8239, IQR = 0.1994) while movements in the Y-axis (Median r= 0.0496, Q1 = -0.1381, Q3 = 0.4331, IQR = 0.5712) were uncorrelated, which is expected given that the arms oscillate out of phase as mice alternate reach and pull epochs to advance the string (Fig. 2c). After iatrogenic injury to the SS/IS tendons, we observed qualitative changes to the shape of the Y-axis string pulling waveform including decreased velocity of pulls as well as “rounding” of the waveform peak at each reach-to-pull transition (Fig. 2d, same mouse as in Fig. 2a). In order to quantify the coordination across the reach-to-pull transition, we calculated the full-width at half maximum (FWHM) of each peak in the waveform (Fig. 2e, example FWHM calculation, *black lines*, shown in the top trace). We also calculated the velocity and acceleration of the arms by taking the first and second derivatives of the Y-axis kinematic trace data, respectively (Fig. 2e, example velocity calculation, *green lines*, shown in bottom trace).

**Figure 2.**
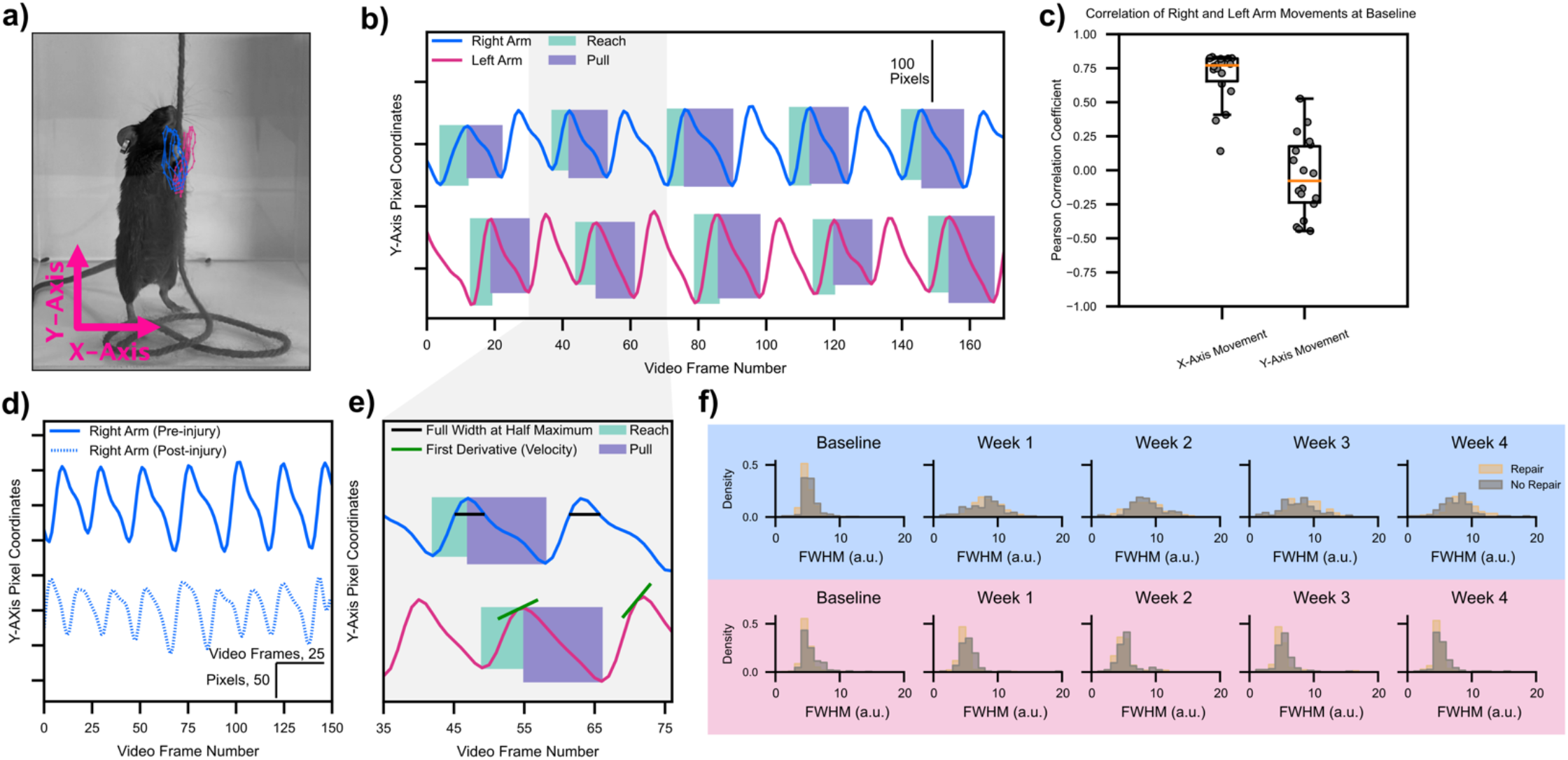
Post-processing kinematic trajectories to extract biomarkers of shoulder function. **a)** Representative mouse with 10 cycles of string pulling overlaid. Right arm in blue, left arm in red. Raw kinematic trajectories with no filtering shown in this example. **b)** Same data as in a) but unrolled across time for the Y-axis. Green boxes represent reach epochs, purple boxes represent pull epochs. Every other epoch labeled for clarity. Amplitude and time are extracted for each reach and pull epoch. Data is highpass and lowpass filtered in b) and all further plots. **c)** Pearson correlation coefficients for X/Y-axis traces, right and left arms pooled. Points represent individual videos from each animal. **d)** Same mouse as in a). Pre-injury Y-axis kinematic trajectory shown as a solid line, post injury trajectory shown as a dashed line. **e)** Expansion of frames 35-75 in b) showing calculation of full width at half maximum (black lines) and velocity (first derivative, green lines). **f)** No difference was noted in quantitative analyses of waveform shape across mice with rotator cuff repair or no repair. Thus mice from both groups were pooled for all further analyses. All statistics are given in Supplementary Table 2.

### Rotator cuff injury impairs movement coordination and dynamic range

We initially were interested in determining if immediate repair of the rotator cuff after iatrogenic injury would accelerate healing. Analyzing the FWHM measure (Fig. 2e) for the right hand (light blue background) and the left hand (light red background) revealed no differences in waveform shape for animals with and without immediate rotator cuff. We thus collapsed the repair and no-repair groups together for all further analysis. Examining the FWHM between the Baseline and Week 1-4 recordings reveals a striking post-injury rightward shift in the values suggesting that animals progress slower across the reach-to-pull transition (Fig. 3a). In contrast to changes in FWHM, the distribution of the velocity values for the injured arm exhibited a smaller relative change in mean (Fig. 3b). Specifically, we observed reduced probability density over negative velocity values, which is consistent with qualitative observations of reduced slope over the pull epoch (Fig. 2d) and suggests that the pull phase is especially affected by decreased slope magnitude. Despite this shift in density, a two-way ANOVA reveals no change in the central tendency of the mean for velocity based on injury, time, or the interaction between both. However, a Levene’s test for equality of variance found statistically significant differences in the variance of the velocity distribution when comparing the uninjured and injured arms at each weekly time point. Similar to velocity, we noticed a drop in the density of negative acceleration values (Fig. 3c). This finding again suggests that, after undergoing rotator cuff injury, mice were unable to generate rapid arm motions, especially in the downward direction. However, in contrast to velocity, where there was no change in central tendency of the mean, acceleration exhibited a significant change in mean value across the experiment with both a main effect of arm (injured vs control) as well as time. Moreover, a Levene’s test for equality of variance found statistically significant differences in the variance of the acceleration distribution when comparing the uninjured and injured arms at each weekly time point. We also found that neither FWHM, velocity, or acceleration recovered across the four post-operative weeks, suggesting lasting deficits in end-effector control secondary to rotator cuff injury.

**Figure 3.**
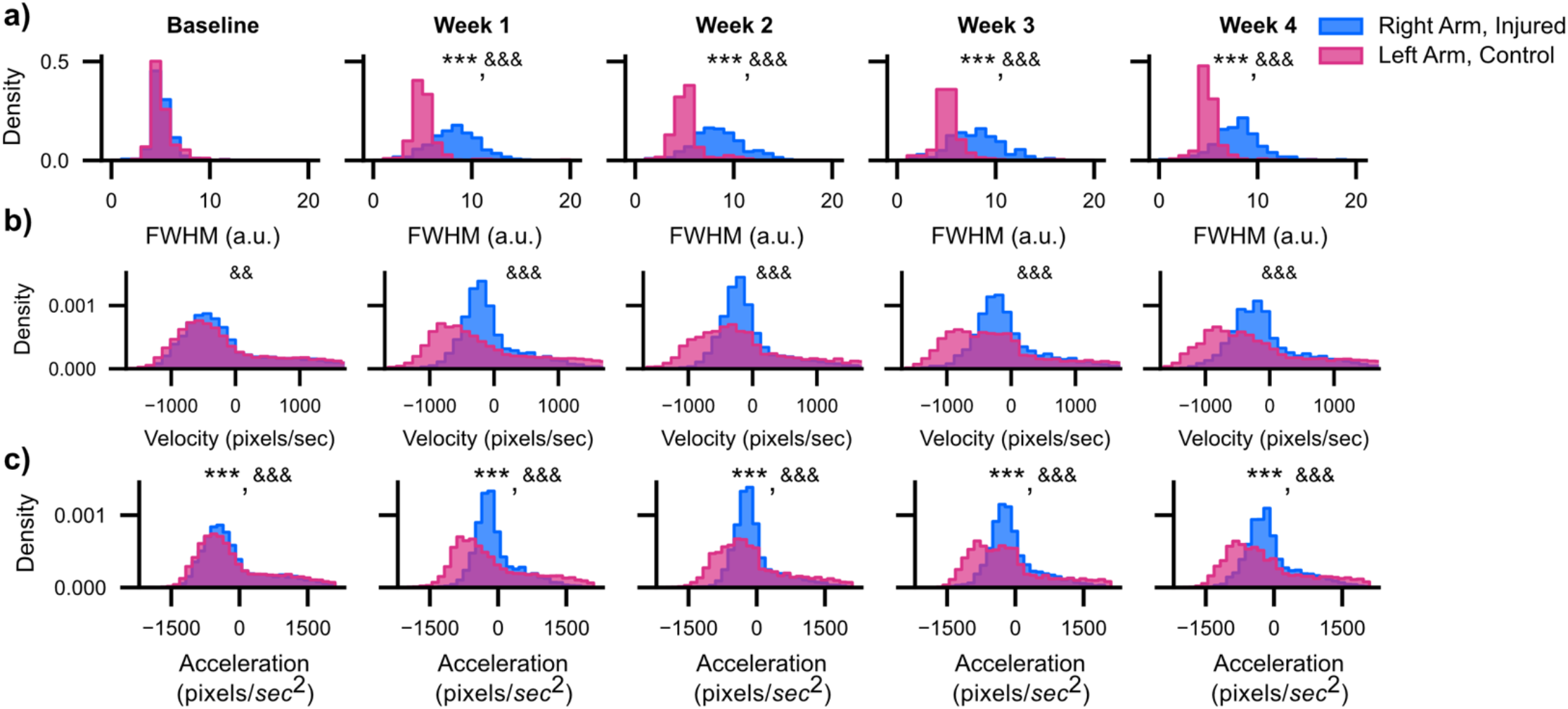
Quantitative measures of waveform shape do not recover after injury. **a)** Histogram of FWHM values for the right (injured) and left (control) arms across all mice and all waveform peaks (n=12 mice). **b)-c)** Same as a) only for velocity and acceleration. *<0.05; **<0.01; ***<0.001, two-way ANOVA, Tukey multiple comparison corrected post-hocs. &<0.05; &&<0.01; &&&<0.001, Levene’s Test for equality of variance. All statistics are given in Supplementary Table 2.

### Kinematic synergies are disrupted by injury and gradually recover across time

Previous work has found that movements of high-dimensional effectors performing everyday tasks can be expressed using a weighted combination of a few kinematic postures^20,21^, or “movement synergies.” Synergies may reflect an innate adaptation of the central nervous system used to simplify coordination of movement across joints with high degrees of freedom^20,22^. Here we used principal component analysis (PCA, implemented using singular value decomposition) to identify covariation patterns in the X-/Y-axis movement traces of the right and left arms into kinematic synergies that are expressed as low dimensional principal components.

After running the PCA algorithm on the X-/Y-axis movement traces of the right and left arms from each video, we next analyzed the cumulative variance explained by PCs 1 through 4. Here, we found that the cumulative variance explained by PCs 1 and 2 across all time points accounted for about 90% of the variability in the data (Supp. Fig 1). Thus, we chose to focus our analyses only on the first two PCs. In general, the higher the variance explained by a smaller number of PCs, the lower dimensional the movement synergy (and the more temporally coordinated the movement across time). In Figure 4a, we show the variance explained by PC1 and PC2 individually. After RC injury, the variance explained by PC1 decreased from its peak at baseline before increasing again at four week’s recovery (Variance explained by PC1, Mean ± SEM, n=12 mice: Baseline 0.707±0.007, Week 1 0.624±0.019, Week 2 0.616±0.029, Week 3 0.600±0.015, Week 4 0.645±0.021).

**Figure 4.**
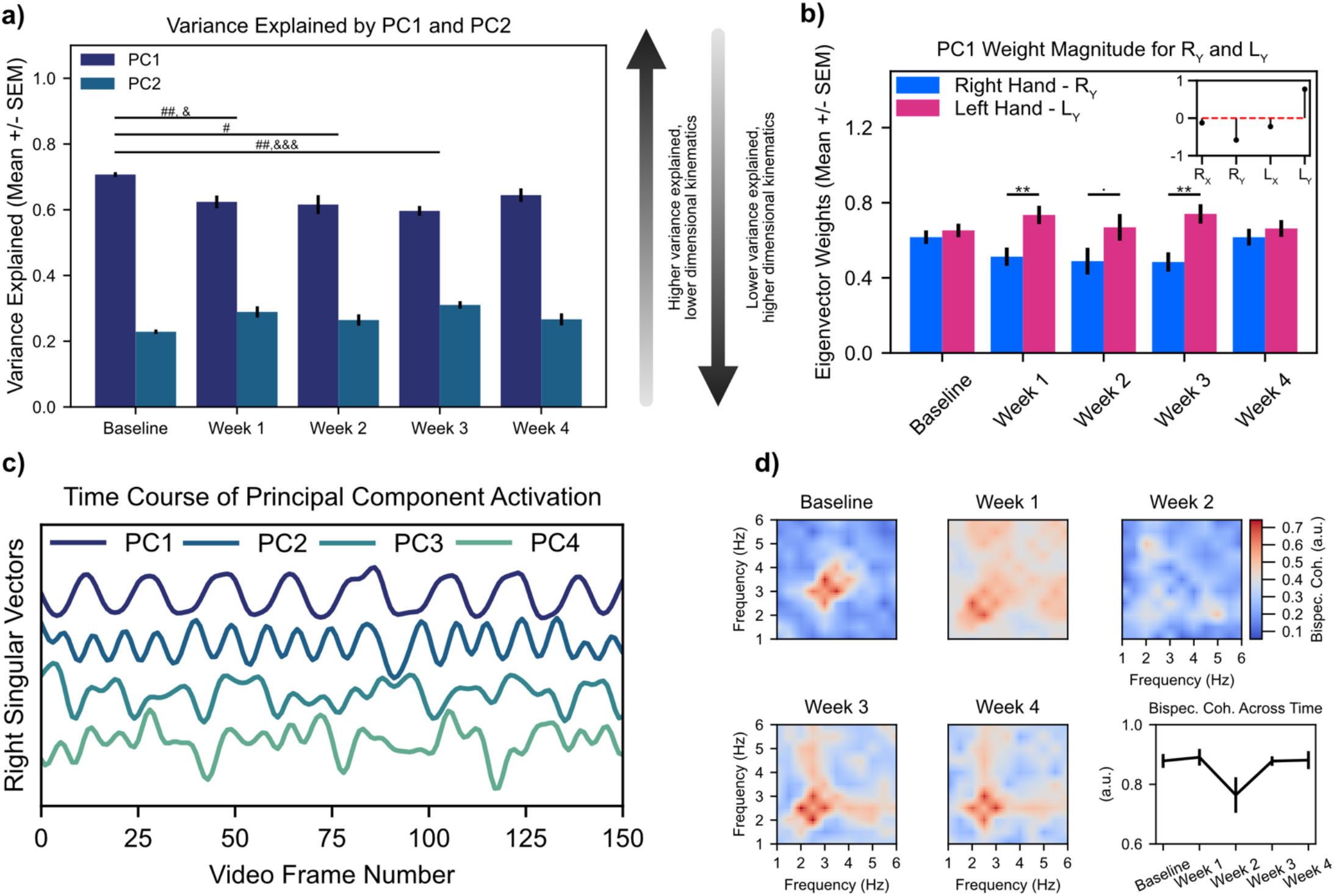
Changes in kinematic synergies track shoulder injury and recovery. **a)** Quantification of variance explained by the first two PCs. #<0.05; ##<0.01; ###<0.001, Tukey multiple comparison corrected post-hocs comparing PC1 variance explained relative to Baseline. &<0.05; &&<0.01; &&&<0.001, Tukey multiple comparison corrected post-hocs comparing PC2 variance explained relative to Baseline. **b)** Quantification of the absolute magnitude of Y-axis Eigenvector weights for right hand (injured) and left hand (uninjured). Inset stem plot shows Eigenvector weights for the video recording from the example mouse in Fig. 2a. ·<0.1, *<0.05; **<0.01; ***<0.001, Tukey multiple comparison corrected post-hocs comparing injured vs uninjured arms. **c)** Representative plot of right singular vectors from a singular value decomposition (i.e., principal component analysis, PCA) of X and Y kinematic trajectories of the right and left arms for same mouse as shown in Fig. 2a. **d)** Mean bispectral coherence cross-coupling between PC1 and PC2; note deterioration of cross-frequency coupling during post-injury weeks 1 and 2 followed by reemergence of coupling during weeks 3 and 4. All statistics are given in Supplementary Table 2.

Meanwhile, the variance explained by PC2 increased from its minimum value at baseline (variance explained by PC2, Mean ± SEM, n=12 mice: Baseline 0.229±0.007, Week 1 0.289±0.017, Week 2 0.265±0.017, Week 3 0.310±0.011, Week 4 0.267±0.018). There was a statistically significant main effect of time on variance explained by PC1 for Baseline versus all weeks, except Week 4. For PC2, there was a statistically significant difference between the variance explained by PC2 for Baseline vs. Weeks 1 and 3 but not for Weeks 2 or 4.

While quantifying the percent variance explained by the first two PCs provides insight into the dimensionality of the kinematic synergy governing movements of the right and left arms, it does not provide insight into how the right and left hands are individually contributing to the synergy. By examining the eigenvectors of the PCA decomposition, we can gain insight into the magnitude and direction with which each of the four kinematic variables (Right arm X-axis (R_X_), Right arm Y-axis (R_Y_), Left arm X-axis (L_X_), and Left arm Y-axis (L_Y_) trajectories) contribute to each PC. The inset plot in Figure 4b shows a stem plot of the eigenvector weights for PC1 for a representative animal at baseline (see Supp. Fig. 2 for plots of absolute eigenvector weights for the other all four PCs). Here, we found that at baseline movements of the right and left arms in the Y-axis were similar in magnitude but opposite in sign—as expected given that the arms oscillate out of phase during the string pulling behavioral cycle. Since the sign of an eigenvector is relative, we took the absolute value of the eigenvector weights for R_Y_ and L_Y_ to compare changes in weights across time (Fig. 4b). At Baseline, both R_Y_ and L_Y_ had similar magnitudes before diverging on Weeks 1-3. This divergence was followed by a convergence of weights on Week 4. Together, these results suggest that after injury the uninjured hand contributes more strongly towards coordinated activity within PC1 as compared to the injured arm; recovery of eigenvector weights on Week 4 further suggests that PCA may be useful in identifying reestablishment of coordinate kinematics during recovery.

In our PCA analysis we have thus far focused on analyzing variance explained and eigenvector weights, which provide insight into the dimensionality of movement coordination as well as the contribution of individual variables to each dimension, respectively. However, neither of these metrics provide temporal information as to patterning of principal component activation. To understand the temporal evolution of each principal component across each video recording, we analyzed the right singular vectors generated as part of the PCA algorithm capture the (Fig. 4c, time series of PC1-4 activation for a representative mouse). In a representative example at baseline, we noticed clear 2-4 Hz coupling between the right singular vectors of PC1 and PC2 (Fig. 4c, *top* and *second from the top* traces)—in other words, for every one oscillatory cycle of PC1, PC2 exhibits two-to-four oscillatory cycles. Thus, to quantify the relationship between the right singular vectors of PCs 1 and 2, we performed a bispectral coherence analysis^23^. Bispectral coherence quantifies cross-frequency coupling between two time series; we found strong coupling between PCs 1 and 2 during the pre-injury baseline (Fig. 4d, mean bispectral coherence values shown across n=12 animals). Following injury, the crisp cross-frequency coupling seen during the Baseline recordings degraded between PC1 and PC2 during post-injury weeks 1 and 2 (Fig 4d, *Week 1* and *Week 2* heatmaps) only to reemerge during post-injury weeks 3 and (Fig. 4d, *Week 3* and *Week 4* heatmaps). To better quantify change in bispectral coherence, we measured the maximal bispectral coherence values in the 2-4 Hz range for each animal and for each video during the recording timeline. Here we found a clear decrease in maximal bispectral coherence values for Week 2. However, a one-way ANOVA with a main effect of time found only a trend towards statistical significance. Together, these results show that injury results in only transient temporal decoupling between PCs 1 and 2, which perhaps suggest rapid central or peripheral adaptation of end effector control to reengage movement synchrony over recovery.

### Rotator cuff injury results in lasting compensation by the contralateral arm

Our waveform shape analysis revealed lasting deficits in the reach-to-pull transition while our kinematic synergy analysis shows that while injury caused the temporary emergence of a higher dimensional movement state space, this dimensionality reverted to baseline at the end of four week’s recovery. How then can we reconcile these two competing findings? Analyzing movement amplitude (Fig. 5a) showed a striking decrease in movement amplitude of the right arm after rotator cuff injury between Baseline and Week 1. Over the ensuing three weeks of recovery (Week 2 through Week 4), movement amplitude for the injured right arm recovered back to its Baseline while movement amplitude for the left arm exceeded that of the pre-injury baseline, suggesting that mice continue compensating with their left arms even after right arm kinematics recovers. This effect was seen symmetrically across both reaching (Fig. 5a, *solid lines*) and pulling (Fig. 5a, *dotted lines*) epochs. A three-way ANOVA with main effects of time, arm (injured versus uninjured), and movement epoch (reach versus pull) plus an interaction term of time and arm was fit to the amplitude data. We found statistically significant main effect of time plus a significant effect of the interaction between time and arm, but there was no significant main effect of arm. As expected, there was no significant effect of movement epoch. A Tukey multiple comparison corrected post-hoc analysis showed statistically significant differences in the interaction term when comparing Baseline relative to post-injury Week 1 for the injured arm, but not for Baseline vs. Weeks 2-4 and for the uninjured arm for Baseline vs. Week 2. These statistical findings further reinforce how, after just one week’s recovery time, the right arm movement amplitude returns back to its pre-injury baseline while the left arm exhibits persistent compensation.

**Figure 5.**
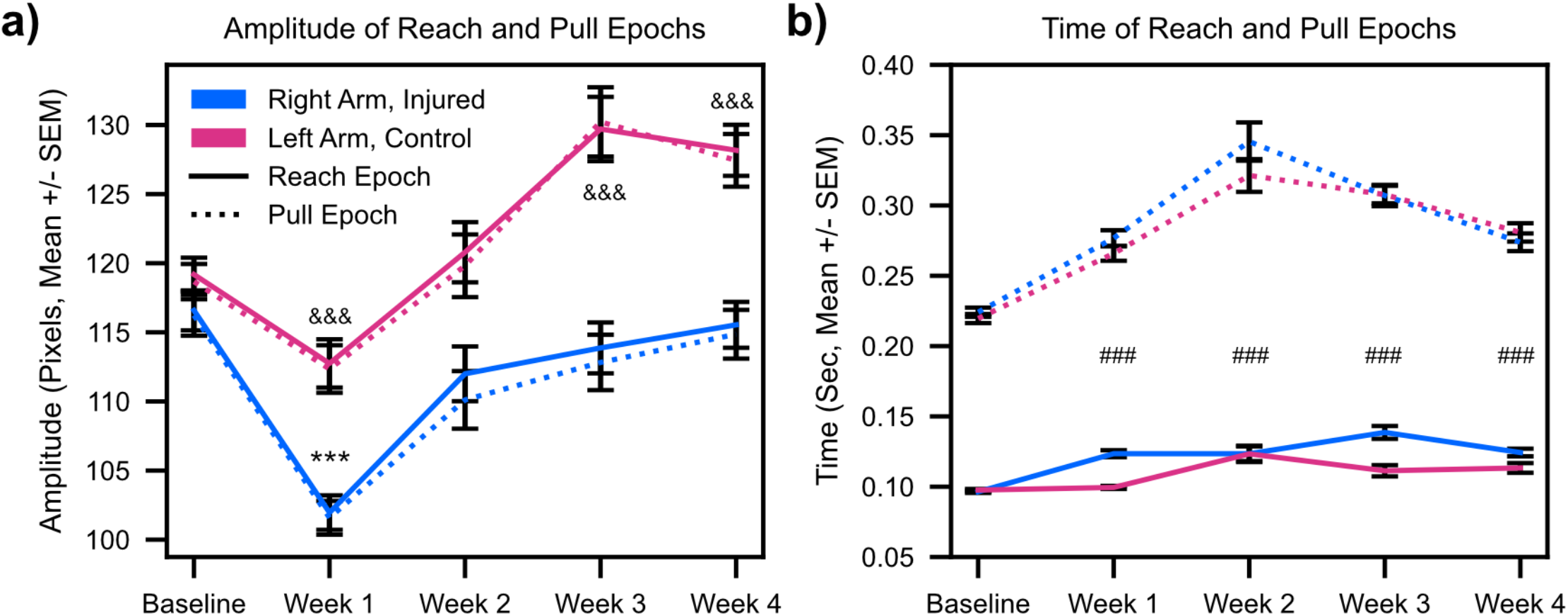
String pulling movement amplitude and time show evidence of compensation. **a)** Amplitude (in pixels) of reach (solid lines) and pull (dashed lines) for right (injured) and left (control) arms. *<0.05; **<0.01; ***<0.001, Tukey multiple comparison corrected post-hocs comparing amplitude for injured arm vs Baseline. &<0.05; &&<0.01; &&&<0.001, Tukey multiple comparison corrected post-hocs comparing amplitude for uninjured arm vs Baseline. **b)** same as a) only for reach and pull times. #<0.05; ##<0.01; ###<0.001, Tukey multiple comparison corrected post-hocs comparing the main effect of time for both arms relative to Baseline. All statistics are given in Supplementary Table 2.

In parallel with quantifying movement amplitude, we also quantified movement time for reach and pull epochs (Fig. 5b). Here, we noticed a clear difference between movement epoch with pulls universally taking longer than reaches (Fig. 5b, pull times shown as *dotted* lines and reach times shown as *solid* lines). At Baseline both reach and pull times are highly symmetrical. This symmetry is followed by an overall trend to longer reach times for the right arm after rotator cuff injury. Pull times also lengthen, albeit more symmetrically, for both the right and left arms before reaching a peak at Week 2 and then declining for the remaining two weeks. We hypothesize that the parallel increase in pull times is attributable to different mechanisms for the injured and uninjured arms—the former because injury slows movement and the latter because of compensation which requires the execution of higher amplitude movements. We fit a three-way ANOVA with main effects of time, arm (injured versus uninjured), and movement epoch (reach versus pull) plus an interaction term of time and arm onto the time data. We found statistically significant main effects of time and movement epoch, plus a trend towards a statistically significant effect of the interaction between time and arm. There was no significant effect of injured vs. uninjured arm. Tukey multiple comparison corrected post-hoc analysis showed statistically significant differences in the main effect of time for Baseline vs. Week 1-4.

### Human patients with rotator cuff injuries recapitulate the kinematic phenotype seen in rodents

Having used the precise temporal control over injury afforded by a rodent model to develop a clear set of measures that differentiate the kinematics of injured vs uninjured shoulders, we next sought to validate our biomarkers on human patients with known RC tears (n=6, see Supp. Table 3 for patient demographic information) as well as healthy controls with no shoulder pathology (n=6). We tracked the position of both the hands and elbows for our human subjects (Fig. 6a, three cycles of string pulling in a representative control subject). Unrolling the kinematic trace in the Y (vertical)-axis across time reveals striking qualitative similarities in waveform shape across rodents (Fig. 2b) and humans (Fig. 6b, same representative control subject as in 6a with the entire video unrolled across time in the Y-axis): reaches exhibit faster rises as compared to pulls, and both arms oscillate out of phase with respect to each other.

**Figure 6.**
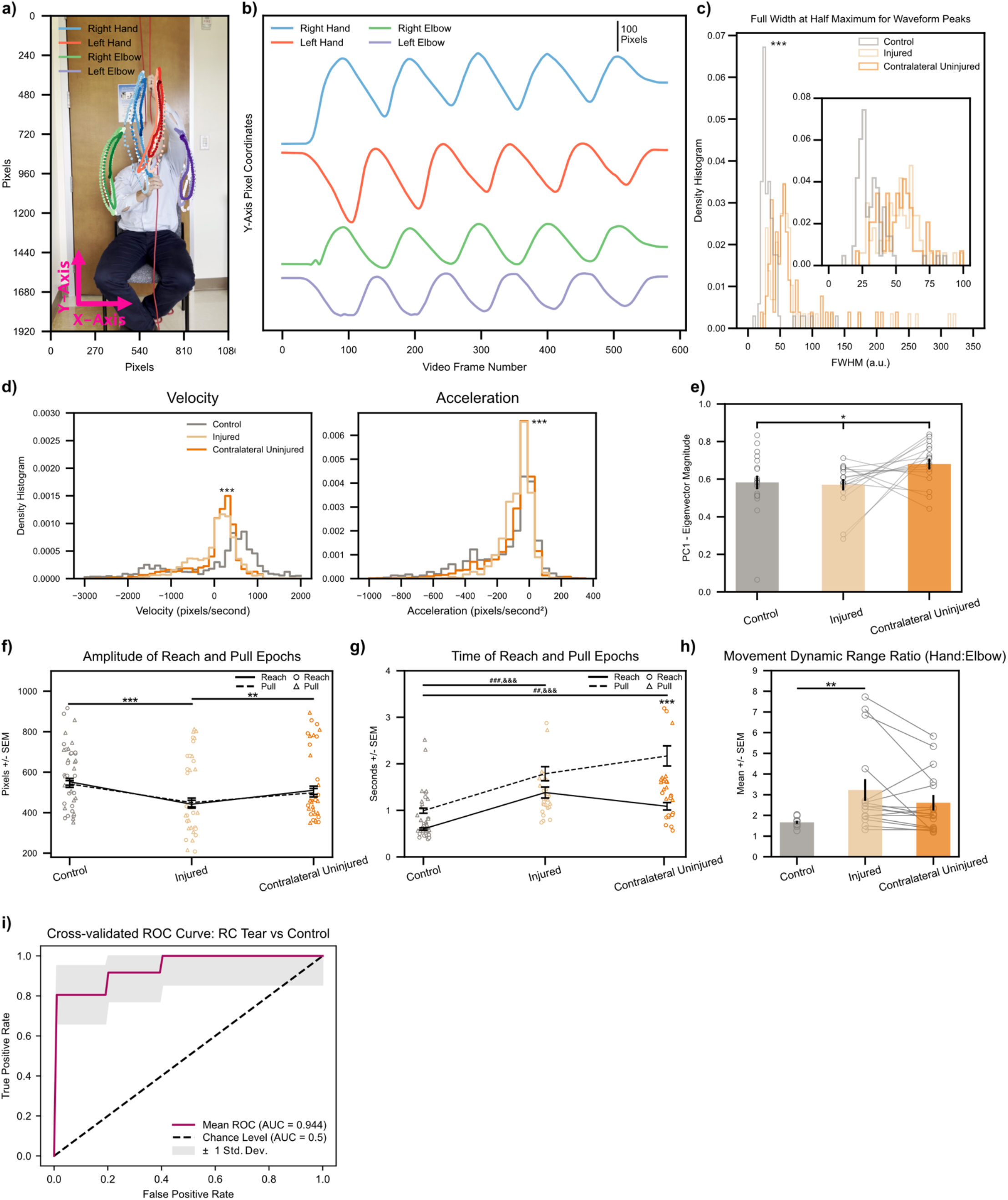
Biomarkers calculated on string pulling kinematic traces translate directly to human subjects with shoulder injury. **a)** Representative control subject with three cycles of string pulling superimposed. Elbows were labeled in addition to hands for the human patients given the ready visibility of human elbows. **b)** Same subject as in a), data shown for one full trial. Note similarity of kinematic trajectories for the hands between human and rodent subjects (Fig. 2b). **c)** FWHM measurements for control (n=12) and injured shoulders (n=6). Inset shows zoomed view for FWHM values between 0 to 100. ***<0.001, Kolmogorov-Smirnoff test. **d)** *Left*, histogram of velocity values (calculated on the Y-axis kinematic trajectories of the hands) for control (n=6), injured (n=6), and contralateral uninjured (n=6) shoulders. *Right*, same as *left* only for acceleration values. ***<0.001, Kruskal-Wallis test. **e)** Quantification of absolute Eigenvector weights for the first PC (data shown for Y-axis Eigenvector weights of the hands). Gray lines show change between injured and contralateral uninjured shoulders for each trial recorded per subject. *<0.1, one-way ANOVA **f-g)** Amplitude and time of reach (solid line) and pull (dashed line) epochs. Individual amplitude/time values for every cycle of string pulling shown for reaches and pulls with circles and triangles, respectively. **h)** Ratio of standard deviation values between ipsilateral hand:elbow pairs calculated for each subject on their hand/elbow Y-axis kinematic trajectories. Gray lines show change between injured and contralateral uninjured shoulders for each trial recorded per subject. *<0.05; **<0.01; ***<0.001, two-way ANOVAs, Tukey multiple comparison corrected post-hocs for all other statistical analyses unless stated otherwise. **i)** Receiver operating characteristic (ROC) curve for a binary logistic regression model fitted to predict a patient as either having no RC tear or having an RC tear in one of their shoulders. *Maroon* line shows mean ROC across 3-fold stratified cross-validation, *gray* outline plots +/-1 Std. Dev. uncertainty in the mean ROC estimate. All statistics are given in Supplementary Table 2.

We next proceeded to validate whether waveform shape quantitatively differs across injured vs uninjured shoulders. As in mice, we used only data for hand movements and found that patients with injured shoulders exhibited increased FWHM values of the Y-axis string pulling waveform peaks (Fig. 6c, data from control shoulders irrespective of laterality in grey. Data from injured shoulders shown in light gold. Inset provides zoomed view on histogram values on the interval from [0, 100]. See Supp. Fig. 4 for FWHM histograms of individual study participants). In parallel with analyzing waveform shape, we also took the first and second derivatives of the waveform trajectories to analyze velocity and acceleration, respectively (Fig. 6d). Just as in rodents, a histogram of velocity (in pixels/second) revealed an increased concentration of values around zero (control: 7.853±28.661) for both the injured shoulder (injured: 2.207±13.970) and, curiously, the contralateral uninjured shoulder (contralateral uninjured: 3.782±17.398) in patients with rotator cuff tears. In fact, a one-way Kruskal-Wallis test found a statistically significant main effect of arm on velocity. A Bonferroni-corrected Mann-Whitney U test as a post-hoc analysis showed statistically significant differences in velocity between Control vs Injured, Control vs. Contralateral Uninjured, and Injured vs. Contralateral Uninjured. When analyzing acceleration values (in pixels/second^2^), we do not observe as a striking of a difference between patients and controls (control: - 153.278 ±6.921, injured: -77.339±2.916, contralateral uninjured: -97.407±4.463). However, a Kruskal-Wallis test still reached statistical significance. Follow-up Bonferroni-corrected Mann-Whitney U test as a post-hoc analysis showed statistically significant differences in velocity between Control vs Injured or Contralateral Uninjured, but not for Injured vs. Contralateral Uninjured.

In contrast to our findings in rodents, PCA decomposition of lowpass filtered, mean-centered X/Y position data of the hands and elbows did not reveal a statistically significant difference in dimensionality between control vs rotator cuff tear groups (Supp. Fig. 5). However, there was a trend towards statistical significance in our analysis of Eigenvector magnitudes (Fig. 6e) for Y-axis movements of the hands across the control shoulder, injured shoulders, and contralateral uninjured shoulders.

When longitudinally testing injured mice on the string pulling task, we notice persistent compensation by the contralateral, uninjured extremity that persists even as the kinematics of the injured extremity recovers (Fig. 5a). Here we analyzed the same metrics of amplitude and time for our human subjects. We found that there was no statistically significant difference in movement amplitude (Fig. 6f) for reach and pull epochs across all control, injured, and contralateral uninjured shoulders nor was there a statistically significant interaction between reach/pull epochs and arm. There was, however, a statistically significant main effect of arm on amplitude. A Tukey multiple comparison corrected post-hoc analysis showed statistically significant differences in the main effect of group for Control vs. Injured or Contralateral Uninjured, but not for Control vs Contralateral Uninjured. Together, these results confirm that patients with rotator cuff injury have reduced movement amplitude of the injured extremity.

When analyzing the timing of reach and pull epochs (Fig. 6g, outlier points >4 seconds removed for clarity. See Supp. Fig. 4 for plot with all data points shown), we notice that pulls (dashed line) generally take longer than reaches—a replication of the phenomenon that we see in our rodent data (Fig. 5b *dashed* lines). Mean time in seconds ± SEM for control group (reach: 0.601 ± 0.026, pull: 0.993 ± 0.055), injured (reach: 1.383 ± 0.118, pull: 1.788 ± 0.152), and contralateral uninjured (reach: 1.087 ± 0.081, pull: 2.169 ± 0.216). A two-way ANOVA revealed a statistically significant main effect of epoch as well arm; the interaction between epoch and group was also statistically significant. Curiously, we notice that the contralateral uninjured arm pull time is greater than the contralateral uninjured arm reach time. The divergence in these two measures is striking given the downward trend in reach times for the contralateral uninjured arm vs both reach and pull times for the injured arm. While difficult to ascertain with certainty, we suspect that this may be a manifestation of compensation where, in order to advance the string by an equivalent distance during reach and pull epochs (Fig. 6f), patients are recruiting scapular or thoracic motions thus prolonging the movement cycle.

We next compared the dynamic range ratio, computed by taking the ratio of standard deviation values of the lowpass filtered, mean-centered Y-axis kinematic trajectory between ipsilateral hand:elbow pairs across all study participants (Fig. 6h). Here we were interested in determining whether rotator cuff injury predisposes patients to adopting a movement regime where the string is advanced by rotating the humerus around its longitudinal axis vs engaging the entire arm in reaching & pulling motion (see Supp. Fig 6 for example kinematic traces of control vs RC tear subject demonstrating reduced elbow excursion). In other words, we expect the dynamic range ratio to increase for patients that predominately advance the string by moving their hands using rotational (rather than translational reaching) movements while keeping the elbow stationary. Indeed, we notice a statistically significant increase in the movement dynamic range ratio. The mean standard deviation ± SEM across groups was 1.665 ± 0.069 for control extremities, 3.224 ± 0.525 for injured extremities, and 2.613 ± 0.373 for contralateral uninjured extremities.

As a final analysis, we used regularized binary logistic regression (see *Materials and Methods* for further details of model design) to build a predictive model for whether a given patient had a rotator cuff tear in at least one of their shoulders. After running stratified K-fold cross-validation, we found that using string pulling metrics as predictors resulted in an area under the curve (AUC) of 0.944 +/-0.051 (Mean +/-Std. Dev.) for our classifier. We further noted a mean sensitivity and specificity of 1.0 and 0.806 averaged across the K-fold cross validations, respectively. Even though our sample sizes are relatively small (n = 6 shoulders with RC tears), these results provide preliminary validation for the clinical utility of our movement assessment methodology.

## Discussion

We found that the string pulling task was a sensitive assay for assessing both human and rodent shoulder function. After injury to the RC, mice exhibited changes in quantitative measures of waveform shape (Figs. 2 and 3) that persisted for the duration of the experiment even as straightforward kinematic measures (such as movement amplitude) were restored back their pre-injury baseline (Fig. 5). We also noticed disruptions in cross-frequency coupling of kinematic synergies across injury and recovery as well as the emergence of a higher dimensional kinematic state space after injury (Fig. 4). All of our pre-clinical biomarkers translated directly to human patients and exhibited similar phenotypic shifts with RC injury (Fig. 6) with the exception of changes in kinematic synergies.

### String pulling as a novel animal model of shoulder function that translates well to humans

Previous animal models of rodent tendon injury and repair have invariably drawn inferences about upper or lower extremity function using quadruped gait tasks. In these models, animal subjects are tasked with walking either on a transparent treadmill or bromophenol blue coated paper and the resultant paw prints are scored for measures such as stride length, stride width, paw print area, paw length, or toe spread^24–28^. More recent techniques have included force sensor measurement as rats walk through a transparent treadmill^29^. While these methods have demonstrated functional differences between subtypes of RC tendon injury^25^ or RC tendon repair strategies^28,29^, no study has ever analyzed forelimb function in rodents using bimanual forelimb movements that are analogous to the types of motions that humans perform in daily life. The string pulling assay has multiple advantages over quadruped gait tasks: 1) it allows for the kinematic assessment of each arm independently (and outside the gait cycle) thus allowing for a within-animal control using the contralateral extremity, 2) the task includes a significant overhead motion component which is frequently impaired in patients with RC tears and that challenges the supraspinatus muscle (one of the most commonly injured muscles in the RC)^3^, 3) movements of the lower extremities are decoupled from movements of the arms, and 4) existing literature, plus our own experimental results, suggest strong kinematic concordance between rodents and humans in string pulling task performance^12^. Moreover, the assay is inexpensive requiring only a piece of string, a transparent box (for rodent experiments), and a smartphone capable of video recording. Image segmentation of the resulting videos using deep convolutional neural networks may now be easily accomplished on computers equipped with consumer-grade Graphical Processing Units (GPUs)^19^.

### Waveform shape and kinematic synergy analyses provide latent insights into movement quality

#### Waveform shape

even though we observed clear differences in string pulling movement speed and amplitude between injured and uninjured extremities in both rodents and humans (Figs. 5 and 6), the oscillatory nature of the behavior allows for application of time series analysis techniques. Here we analyzed the full width at half maximum (FWHM) for waveform peaks (i.e., capturing the reach-to-pull transition) of Y-axis string pulling kinematic trajectories. We focused our analysis on the reach-to-pull transition as it 1) captures overhead reaching movements that require the supraspinatus muscle and 2) it highlights the transition from a concentric to an eccentric RC muscle contraction regime. Importantly, FWHM is not simply co-linear with movement duration: in Fig. 5b we notice that both injured and uninjured arms have prolonged pull durations after injury yet the distribution of FWHM undergoes a rightward shift only for those animals with RC injury (Fig. 3a). While further research into this phenomenon is required, we suspect that FWHM may serve as a surrogate measure for neuromuscular control of the reach-to-pull transition. In animals and humans with RC injury, altered proprioceptive input from the injured tendon, or nociceptive input from the joint itself, might necessitate increased cognitive control over the shoulder’s musculature from the central nervous system to switch movement regimes. Alternatively, muscle inhibition secondary to pain may manifest as selective deceleration during the reach-to-pull transition.

#### Kinematic synergies

Prior human research suggests that the temporal evolution of complex movements across joints with high degrees of freedom (hDOF) such as the hand occurs via the coordinated activation of a low-dimensional kinematic basis set^20,30,31^. Analogous to the hand, the shoulder joint occupies a high dimensional anatomic space with 8 degrees of motion and 18 different muscles controlling its articulation. Intriguingly, for human string pulling behavior (where we have access to an eight-dimensional kinematic space from the X/Y position data for four arm markers), only two principal components are sufficient to explain >90% of the variance in the data (see Supp. Fig 5). This suggests that despite the requirement placed upon the body to efficiently covary shoulder flexors, extensors, and stabilizers to generate a smooth, sinusoidal movement, string pulling behavior manifests itself as a predominately low-dimensional activity. Moreover, to our knowledge no prior research has been done studying the effects of musculoskeletal injury and recovery on kinematic synergies across shoulders. In rodents, where we have ready access to longitudinal data, we notice an increase in the dimensionality of the kinematic state-space after injury which recovers back to baseline at the end of four weeks’ recovery. The cross-frequency coupling between PC1 and PC2 undergoes a similar pattern. Curiously, we saw no statistically significant differences in the dimensionality of string pulling behavior in humans with or without RC tears; future studies are necessary to determine if changes in movement dimensionality are a phenomenon of early injury or are linked to specific RC tendon injury patterns. While we do not have access to neural or electromyographic (EMG) data, our work provides a robust behavioral assay for probing these substrates in future studies. Furthermore, the presence of clear biomarkers between injured and non-injured shoulders provides an inexpensive and straightforward means of testing different therapeutics aimed to improve RC function.

### Empowering computerized physical therapy at scale

Here we have developed an inexpensive assay for assessing shoulder function across species. Both the mouse and human video recordings were collected using a smartphone. Given the rise of matrix math “coprocessor” chips embedded into recent-generation smartphone devices, it is now possible to perform image segmentation and analysis entirely on a smartphone device without uploading video over the internet. By deploying our metrics calculated from the string pulling task through a user-facing smartphone app, we can provide privacy-preserving measurements of shoulder health to the community. This new paradigm allows patients with less access to in-person care, especially those from underrepresented backgrounds or without reliable or fast transportation to clinics (e.g. the elderly or those living in rural areas), to easily receive quick and affordable snapshots of their shoulder health using tools they are likely to already have at home. We further envision that patients could use such measures to track their recovery from shoulder injury or surgery. Indeed, one can imagine a future system where our metrics calculated on sinusoidal signals derived from simple physical tasks can be used to diagnose MSK injuries in other areas, such as the back, hip, or knee as well as track functional recovery using easily detected biomarkers without ever having a patient leave their home.

## Materials and Methods

### Animal training protocol and behavioral box apparatus

All animal procedures were approved by the SFVA IACUC committee. 12 adult male wild-type mice (C57/L6J, Jackson Laboratory inc.) were trained on a string-pulling task in an acrylic box as described by Blackwell et. al^11^. After 2 weeks of training, the mice were split into two surgical groups. Mice were placed in a plexiglass box with dimensions (5 by 6 by 9 inches) for training and video recording of string pulling behavior. 3D printed string holders were used to standardize string placement in each box.

### Iatrogenic rotator cuff injury

One group (n=6) underwent a right supraspinatus (SS) and infraspinatus (IS) tendon transection and denervation (TTDN) while another group (n=6) underwent right SS and IS TTDN with immediate repair of the torn tendons as described by Wang et al^28^. String pulling behavior was recorded for all mice at their preoperative baseline and at postoperative weeks 1, 2, 3 and 4. Prior to each recording on the postoperative weeks 1-4, mice were given a brief string pulling training session as a reminder of the task requirements.

### Video Recordings

A 1920×1080 HD video camera recording at 59.94 frames-per-second was used to acquire string pulling videos. The location of the camera relative to the plexiglass box was fixed across sessions using an alignment jig. Video recordings (which were ∼15-30 seconds in length for each pulling trial) were then trimmed with *ffmpeg* v4.4.2 to contain only the string pulling behavior.

### Kinematic Segmentation

The X/Y coordinates of each hand were acquired with *DeepLabCut* (DLC) v2.2.0 using a ResNet50 deep convolutional neural network model. Two DLC models were built across the experiment: 1) a pilot cohort of 3 mice and 2) a full model that included data from all 13 mice pooled together (including mice in the pilot experiment). The locations of the right and left hands were labeled for 320 and 1080 video frames across the pilot and full models, respectively. After initial training, an extra 160 and 1440 frames were extracted as outliers (based on a criteria of ≥ 10 pixel jumps in Euclidean distance between consecutively labeled points across video frames) for the pilot and full models, respectively. Each iteration of the ResNet50 model was trained for 200,000 iterations and the resultant mean Euclidean error in label location (determined on a held-out set of test images consisting of 5% of the frames in each training set) was calculated using the built-in DLC function *evaluate_network* (data shown in F1Civ). A total of 27 videos were used for neural network training; all training was performed using an NVIDIA 2080 Ti GPU with default image augmentation enabled.

### String Pulling Trajectory Trace Post-Processing

Once the X/Y coordinates of each hand were segmented using DLC, the resulting traces were highpass filtered with a first-order 0.75Hz Butterworth filter (to remove trajectory drift from minor postural changes across the pulling cycle) and then lowpass filtered with a third-order 9Hz Butterworth filter (to remove occasional jitter in trajectory segmentation). All subsequent analyses are performed using filtered data. After filtration, the peaks/troughs in the Y-axis (vertical direction) hand trajectory trace were labeled using SciPy’s *find_peaks* function.

### Calculation of Amplitude and Time for Reach/Pull Epochs

Reach epochs were defined as the time between each trough and its successive peak in the Y-axis kinematics trajectory; pull epochs were defined as the time between each peak and its successive trough in the Y-axis kinematics trajectory. For every reach/pull, we measured the amplitude (in pixels) between each successive trough-to-peak and peak-to-trough epoch corresponding to every reach and pull, respectively. Reach/pull time was measured in number of videos frames, divided by the video framerate, between each successive trough-to-peak and peak-to-trough epoch corresponding to every reach and pull, respectively.

### Calculation of the Full Width at Half Maximum

In order to quantify the shape of the string pulling waveform across the experimental timeline, the full width at half maximum (FWHM) was calculated as a hybrid measure of movement fluency during the period of behavior covering both reaching and pulling. To calculate the FWHM, the Y-axis kinematics trace for the right and left hands was mean-centered and the periods of the pulling trajectory between the signals’ negative-to-positive and positive-to-negative zero-crossing was extracted for analysis. Each epoch was interpolated using a 100 point 2^nd^-degree univariate spline, and the FWHM was calculated as the width (in fractional video frame number) of each peak at 1/2 its vertical amplitude. See Fig. 2E for example FWHM values (black lines) overlaid on a representative string pulling waveform.

### Calculation of Velocity and Acceleration

To calculate velocity and acceleration of the right and left hands across the transition from reaching to pulling, the Y-axis kinematics trace for the right and left hands was processed as described in the section on calculating the full width at half maximum of the signal. After extracting the interpolated signal, the first and second derivates were taken as measures of velocity and acceleration, respectively. See Fig. 2E for example velocity values (green lines) overlaid on a representative string pulling waveform.

### Correlation in hand movement

To measure the consistency of string pulling behavior across the right and left hands, we calculated Pearson’s correlation coefficients by correlating X_right_ with X_left_ and Y_right_ with Y_left_ kinematic traces (all correlations were run after data was high pass and low pass filtered). All correlations were run within animal and within day.

### Quantifying Kinematic Synergies Using Principal Component Analysis

The shoulder is a complex joint that allows for multiplanar motion across arm flexion, extension, abduction, adduction, internal rotation, and external rotation. Moreover, shoulder motion is intimately tied to scapular and thoracic mobility as both contribute to stabilization of the upper extremity across its full range of motion; in total, about 20 skeletal muscles contribute to shoulder motion^4^. Prior research on the human hand, has shown significant biomechanical and temporal linking across joints during various hand movements thus suggesting that the biomechanical and neural representations of the hand are significantly lower dimensional than the degrees of freedom conferred by individual muscles and joints would imply^20,21^.

Here we studied kinematic synergies by performing singular value decomposition (SVD) independently on each string pulling epoch from individual mice across weeks. In brief, the filtered kinematics trace containing data for the right hand X-axis movement, right hand y-axis movement, left hand x-axis movement, and left hand y-axis movement were mean-centered and then concatenated into a matrix *T* ∈ *R*^4 *x t*^ with t representing the number of video frames in each video recorded from a given mouse on a given week.

The matrix *T* was decomposed using SVD into the matrices *U* ∈ *R*^4 *x* 4^, *S* ∈ *R*^4 *x*4^, and *V*^*T*^ ∈ *R*^4 *x t*^. The columns (i.e., principal components) of *U* capture the covariation patterns between the four tracked kinematic variables, with the individual weights of each column capturing both the magnitude and direction with which each kinematic variable contributes to that particular component. The absolute value of the weights in the first principal component were used to capture the magnitude of these kinematic variables for right and left hand Y-axis movement in Fig. 4E. The percent variance explained by each principal component is calculated by squaring the singular values in matrix *S* and then dividing each squared singular value by the sum of all squared singular values (reported in Fig. 4D for the first two PCs). The variance explained by each PC has previously been shown to correlate with the dimensionality of the kinematic synergies, with lower variances explained by each individual PC corresponding to a higher dimensional kinematic synergy as more PCs are required to reach the same cumulative proportion of explained variance. Lastly, each row of the matrix *V*^*T*^ captures the relative temporal contribution of each principal component across each video recording.

### Bispectral Coherence (Bicoherence) Analysis

In order to understand the temporal relationship between activation of PC1 and PC2, we used bicoherence to measure the cross-frequency coupling between the right singular vectors corresponding to PCs 1 and 2. In brief, we used the *scipy* ‘spectrogram’ function to take the time-frequency decomposition of right singular vectors of PCs 1 and 2 using an FFT window length of 2 seconds with 1 second of overlap. The bicoherence analysis was then performed as described elsewhere^23^. Because individual videos contained behavioral epochs of differing lengths, the resulting bicoherence values were linearly interpolated between 0-15Hz in .1Hz for each subject.

### Human data recordings

All patients gave their informed consent to participate in the study protocol as approved by the UCSF Committee on Human Research and all procedures were approved by the UCSF Institutional Review Board (IRB). Control and rotator cuff injury patients were recruited through convenience sampling at the UCSF Orthopedic Institute. Study participants were given minimal instruction on how to perform the string pulling task by the lead study author (D.D.) and then allowed to string pull at their own preferred rhythm and kinematic preference. Video was recorded using a tripod-mounted smartphone camera (iPhone 13 Pro Max) set two meters away from each subject. Video was recorded in HD resolution (1920×1080) at 59.94 frames per second. The resulting videos were subsequently processed in DeepLabCut using 968 training frames and two successive refinement steps (outlier frames were defined as those frames with a Euclidean distance between two successively labeled points of ≥ 20 pixels). We labeled both the hands and the elbows for the human recordings as the elbows were readily visible in our subjects vs. rodents where the elbows are hidden by a layer of fur. For subjects in the injury cohort, the arm with the rotator cuff tear was labeled as in the “injured” extremity while the contralateral arm was labeled as the “uninjured extremity.”

### Human data pre-processing

In order to ensure best performance for the detection of peaks/troughs in our analysis of string pulling amplitude & time, the Y-axis kinematic trajectory for the hands was highpass-filtered at 0.1 Hz with a 1^st^ order Butterworth filter followed by a 7^th^ order Butterworth 7 Hz lowpass filter. The peaks/troughs of this signal were than extracted and analyzed analogously as for rodents, described in the section “Calculation of Amplitude and Time for Reach/Pull Epochs” above. For all other analyses (including calculation of the full width at half maximum, velocity, acceleration, and PCA decomposition) we did not highpass filter the X/Y kinematic trajectories for the hands and elbows instead only performing lowpass filtering as the human data was less noisy when compared to the rodent data. Methods for calculating the full width at half maximum, velocity, acceleration, and PCA decomposition of the signals were performed analogously to our methods used for analyzing rodent data (see sections “Calculation of the Full Width at Half Maximum”, “Calculation of the Velocity and Acceleration”, and “Quantifying Kinematic Synergies Using Principal Component Analysis” above).

### Movement Dynamic Range Ratio

Because we are able to track the position of both the hands and the elbows in our human subjects, we calculated a Movement Dynamic Range Ratio that quantified the relative contribution of hand vs elbow movements in the string pulling behavior. The Movement Dynamic Range Ratio was calculated by first taking the standard deviation of the lowpass filtered Y-axis kinematic trajectories for each subject’s hands and elbows. We then took the within-subject ipsilateral ratio between the two standard deviation values (e.g.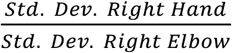) across the right/left hand and elbow. These values were then reported as mean +/-SEM in Fig. 6h. As intuition, if subjects predominantly moved the arm from the shoulder as the main pivot point, the elbows and the hands would exhibit roughly the same vertical displacement in space (i.e. the Movement Dynamic Range Ratio would be close to 1; see Fig. 6h, control shoulders). On the other hand, if subjects immobilize the shoulder and instead move the arm through rotation of the humerus around its longitudinal axis, we would expect a the vertical displacement of the hands to exceed the vertical displacement of the elbows (i.e. the Movement Dynamic Range Ratio would be greater than 1; see Fig. 6h, injured shoulders).

### Logistic Regression Classifier

In order to determine if the biomarkers calculated from the string pulling waveform can serve as predictors for shoulder injury, we built a binary logistic classifier using the *LogisticRegression* class from the *scikit-learn* package. For the right and left arm of each participant’s video we included as independent variables: mean reach time, mean pull time, mean reach amplitude, mean pull amplitude, mean FWHM, mean velocity, symmetry ratio (right:left arm) for mean reach amplitude and time, symmetry ratio (right:left arm) for mean pull amplitude and time, and the dynamic range ratio. The optimal L2 norm regularization parameter (*C*, 0.001) was determined via K-fold cross-validation using the *GridSearchCV* method. In order to calculate the receiver operator characteristic (ROC) curve and the area under the curve (AUC), we used *scikit-learn*’s stratified K-fold cross-validation helper function *StratifiedKFold* (n =3 folds), and all models were fit using the *saga* solver.

### Statistics

Linear mixed-effects (LME) models were used to test the significance of differences in means across weeks and rotator cuff repair status. Using these models accounts for the fact that kinematics from the same animal are more correlated than those from different animals; thus, it is more stringent than computing statistical significance over all subjects^32^. LME models were fit using *statsmodels* v0.13.2 and included a random intercept for each mouse. Levene’s was used to test for differences in variance. All analysis were performed using Python v3.10.5.

## Data Availability

The data and custom code that support the rodent findings in this study are available from the Senior Author (Brian T. Feeley) upon reasonable request. Human subjects data is not available for distribution because study participants did not consent to such sharing.

## Supplementary Material Legends

Supplementary Table 1: Cost of materials for assessing shoulder function in both rodents and humans using the string pulling task.

Supplementary Table 2: Table of statistical

Supplementary Table 3: Human control and rotator cuff patient demographic information.

Supplementary Movie 1: Full-speed representative video recording of string pulling behavior from a single mouse. Same mouse and video as demonstrated in example traces shown in Figure 2 Panels a,b,d, and e.

Supplementary Movie 2: 0.5x speed representative video recording of string pulling behavior from a single mouse. Same mouse and video as in Supplementary Movie 1.

Supplementary Figure 1: Cumulative variance explained by all principal components for all five weeks

Supplementary Figure 2: Absolute eigenvector weights for all principal components.

Supplementary Figure 3: Symmetry measure for amplitude (orange lines) and time (yellow lines). Data shown as ratio of right:left hand values calculated for every video by dividing mean amplitude/time for all reach/pull epochs.

Supplementary Figure 4: Plot of full-width at half maximum with single subject replicates shown as *dotted* lines in the inset.

Supplementary Figure 5: Variance explained by all eight principal components in the human data, split by control vs injured shoulders.

Supplementary Figure 6: Plot of reach and pull times across control, injured, and uninjured shoulders. Same data as main Fig. 6g only with four outlier points shown.

Supplementary Figure 7: Representative traces for control subject (*solid* lines) and a subject with known rotator cuff tear (*dashed* lines). Note loss of elbow motion (*purple* line) relative to hand motion (*red* line) for the subject with a known rotator cuff tear.

## Author Contributions

Conceptualization: D.M.D., M.R.D., X.L. and B.T.F.; Data curation: D.M.D. and D.A.H.; Formal analysis: D.M.D.; Funding acquisition: M.R.D., X.L. and B.T.F.; Investigation: D.M.D., D.A.H. and F.A.G.; Methodology: D.M.D.; Project administration: D.M.D., X.L. and B.T.F.; Resources: D.M.D., X.L. and B.T.F.; Software: D.M.D.; Supervision: D.M.D., X.L. and B.T.F.; Validation: D.M.D.; Visualization: D.M.D.; Writing – original draft: D.M.D. and D.A.H.; Writing - review & editing: D.M.D., D.A.H., X.L. and B.T.F.;

## Acknowledgements

We express our sincere appreciation to Sierra K. Lear for copy-editing and feedback on this work, as well as to Nikhilesh Natraj and Preeya Khanna for input on computational methodology. This work is supported by NIH 1 F31 NS127514-01 to D.D. and NIH 1R01AR072669, VA BLRD 1IO1BX002680 grants to B.T.F. and X.L.

## REFERENCES

1. Tempelhof, S., Rupp, S. & Seil, R. Age-related prevalence of rotator cuff tears in asymptomatic shoulders. J. Shoulder Elbow Surg. 8, 296–299 (1999).

2. Minagawa, H. et al. Prevalence of symptomatic and asymptomatic rotator cuff tears in the general population: From mass-screening in one village. J. Orthop. 10, 8–12 (2013).

3. Lansdown, D. A. & Feeley, B. T. Evaluation and Treatment of Rotator Cuff Tears. Phys. Sportsmed. 40, 73–86 (2012).

4. Terry, G. C. & Chopp, T. M. Functional Anatomy of the Shoulder. J. Athl. Train. 35, 248–255 (2000).

5. Symptoms Matter—Leading Causes of Disability. NCCIH https://www.nccih.nih.gov/about/symptoms-matterleading-causes-of-disability.

6. Freburger, J. K. & Holmes, G. M. Physical Therapy Use by Community-Based Older People. Phys. Ther. 85, 19–33 (2005).

7. Arnadottir, S. A. & Jonsson, B. G. Outpatient physical therapy population has been aging faster than the general population: a total population register-based study. BMC Health Serv. Res. 21, 708 (2021).

8. Urwin, M. et al. Estimating the burden of musculoskeletal disorders in the community: the comparative prevalence of symptoms at diVerent anatomical sites, and the relation to social deprivation. 7 (1998).

9. Liu, X. et al. A Mouse Model of Massive Rotator Cuff Tears. JBJS 94, e41 (2012).

10. Liu, X. et al. Investigating the cellular origin of rotator cuff muscle fatty infiltration and fibrosis after injury. Muscles Ligaments Tendons J. 6, 6–15 (2016).

11. Blackwell, A. A., Banovetz, M. T., Qandeel Whishaw, I. Q. & Wallace, D. G. The structure of arm and hand movements in a spontaneous and food rewarded on-line string-pulling task by the mouse. Behav. Brain Res. 345, 49–58 (2018).

12. Singh, S. et al. Human string-pulling with and without a string: movement, sensory control, and memory. Exp. Brain Res. 237, 3431–3447 (2019).

13. Schwartz, C., Hazée, A., Denoël, V. & Brüls, O. SHOULDER INJURY PREVENTION IN SPORTS USING 3D MOTION CAPTURE. 1.

14. Rawashdeh, S. A., Rafeldt, D. A., Uhl, T. L. & Lumpp, J. E. Wearable motion capture unit for shoulder injury prevention. in 2015 IEEE 12th International Conference on Wearable and Implantable Body Sensor Networks (BSN) 1–6 (IEEE, 2015). doi:10.1109/BSN.2015.7299417.

15. Gritsenko, V. et al. Feasibility of Using Low-Cost Motion Capture for Automated Screening of Shoulder Motion Limitation after Breast Cancer Surgery. PLOS ONE 10, e0128809 (2015).

16. Park, C. et al. Comparative accuracy of a shoulder range motion measurement sensor and Vicon 3D motion capture for shoulder abduction in frozen shoulder. Technol. Health Care 30, 251–257 (2022).

17. Jackson, M., Michaud, B., Tétreault, P. & Begon, M. Improvements in measuring shoulder joint kinematics. J. Biomech. 45, 2180–2183 (2012).

18. Charbonnier, C., Chagué, S., Kolo, F. C., Chow, J. C. K. & Lädermann, A. A patient-specific measurement technique to model shoulder joint kinematics. Orthop. Traumatol. Surg. Res. 100, 715–719 (2014).

19. Mathis, A. et al. DeepLabCut: markerless pose estimation of user-defined body parts with deep learning. Nat. Neurosci. 21, 1281–1289 (2018).

20. Natraj, N., Silversmith, D. B., Chang, E. F. & Ganguly, K. Compartmentalized dynamics within a common multi-area mesoscale manifold represent a repertoire of human hand movements. Neuron 110, 154–174.e12 (2022).

21. Ejaz, N., Hamada, M. & Diedrichsen, J. Hand use predicts the structure of representations in sensorimotor cortex. Nat. Neurosci. 18, 1034–1040 (2015).

22. Safavynia, S., Torres-Oviedo, G. & Ting, L. Muscle Synergies: Implications for Clinical Evaluation and Rehabilitation of Movement. Top. Spinal Cord Inj. Rehabil. 17, 16–24 (2011).

23. Understanding phase-amplitude coupling from bispectral analysis.17.

24. Soslowsky, L. J., Carpenter, J. E., DeBano, C. M., Banerji, I. & Moalli, M. R. Development and use of an animal model for investigations on rotator cuff disease. J. Shoulder Elbow Surg. 5, 383–392 (1996).

25. Perry, S. M., Getz, C. L. & Soslowsky, L. J. Alterations in function after rotator cuff tears in an animal model. J. Shoulder Elbow Surg. 18, 296–304 (2009).

26. Messner, K., Wei, Y., Andersson, B., Gillquist, J. & Räsänen, T. Rat Model of Achilles Tendon Disorder. Cells Tissues Organs 165, 30–39 (1999).

27. Fu, S.-C., Chan, K.-M., Chan, L.-S., Fong, D. T.-P. & Lui, P.-Y. P. The use of motion analysis to measure pain-related behaviour in a rat model of degenerative tendon injuries. J. Neurosci. Methods 179, 309–318 (2009).

28. Wang, Z. et al. A Mouse Model of Delayed Rotator Cuff Repair Results in Persistent Muscle Atrophy and Fatty Infiltration. Am. J. Sports Med. 46, 2981–2989 (2018).

29. Sarver, J. J., Dishowitz, M. I., Kim, S.-Y. & Soslowsky, L. J. Transient Decreases in Forelimb Gait and Ground Reaction Forces Following Rotator Cuff Injury and Repair in a Rat Model. J. Biomech. 43, 778–782 (2010).

30. Bizzi, E., Cheung, V. C. K., d’Avella, A., Saltiel, P. & Tresch, M. Combining modules for movement. Brain Res. Rev. 57, 125–133 (2008).

31. Santello, M., Flanders, M. & Soechting, J. F. Postural Hand Synergies for Tool Use. J. Neurosci. 18, 10105–10115 (1998).

32. Aarts, E., Verhage, M., Veenvliet, J. V., Dolan, C. V. & van der Sluis, S. A solution to dependency: using multilevel analysis to accommodate nested data. Nat. Neurosci. 17, 491–496 (2014).

